# Regular universal screening for SARS-CoV-2 infection may not allow reopening of society after controlling a pandemic wave

**DOI:** 10.1101/2020.11.18.20233122

**Authors:** MCJ Bootsma, ME Kretzschmar, G Rozhnova, JAP Heesterbeek, JAJW Kluytmans, MJM Bonten

## Abstract

**Background:** To limit societal and economic costs of lockdown measures, public health strategies are needed that control the spread of SARS-CoV-2 and simultaneously allow lifting of disruptive measures. Regular universal random screening of large proportions of the population regardless of symptoms has been proposed as a possible control strategy.

**Methods:** We developed a mathematical model that includes test sensitivity depending on infectiousness for PCR-based and antigen-based tests, and different levels of onward transmission for testing and non-testing parts of the population. Only testing individuals participate in high-risk transmission events, allowing more transmission in case of unnoticed infection. We calculated the required testing interval and coverage to bring the effective reproduction number due to universal random testing (*R*_*rt*_) below 1, for different scenarios of risk behavior of testing and non-testing individuals.

**Findings:** With *R*_0_ = 2.5, lifting all control measures for tested subjects with negative test results would require 100% of the population being tested every three days with a rapid test method with similar sensitivity as PCR-based tests. With remaining measures in place reflecting *R*_*e*_ = 1.3, 80% of the population would need to be tested once a week to bring *R*_*rt*_ below 1. With lower proportions tested and with lower test sensitivity, testing frequency should increase further to bring *R*_*rt*_ below 1. With similar *R*_*e*_ values for tested and non-tested subjects, and with tested subjects not allowed to engage in higher risk events, at least 80% of the populations needs to test every five days to bring *R*_*rt*_ below. The impact of the test-sensitivity on the reproduction number is far less than the frequency of testing.

**Interpretation:** Regular universal random screening followed by isolation of infectious individuals is not a viable strategy to reopen society after controlling a pandemic wave of SARS-CoV-2. More targeted screening approaches are needed to better use rapid testing such that it can effectively complement other control measures.

**Funding:** RECOVER (H2020-101003589) (MJMB), ZonMw project 10430022010001 (MK, HH), FCT project 131_596787873 (GR). ZonMw project 91216062 (MK)

## Introduction

Intensive public health measures can effectively control transmission of SARS-CoV-2, yet at high societal and economic costs. Innovative strategies that allow lifting of societally disruptive control measures without increasing transmission and initiating a next pandemic wave are, therefore, urgently needed. Testing for infection with SARS-CoV-2 followed by contact tracing is instrumental in controlling spread, but is seriously hampered by diagnostic delays inherent to the use of high-throughput RT-PCR [Kretzschmar et al. (2020)]. Novel rapid testing technologies, such as antigen-based tests and LAMP, can reduce diagnostic delays to several minutes [Rödel et al. (2020)]. Moreover, such tests can identify symptomatic and asymptomatically infected subjects with high Yet, compared to PCR-based testing, test sensitivity of rapid tests ranges from 70% to 90%, with false-negative results reported mainly in samples with low viral loads (i.e., high CT values), that may reflect subjects with low likelihood of being infectious [Gremmels et al. (2020)]. Nonetheless, such tests can facilitate a shift from finding infected individuals to finding infectious individuals [Mina et al. (2020)], thus potentially having sufficient impact on transmission to control spread effectively, i.e., with a minimum set of acceptable additional measures.

Over the course of the first wave of the COVID-19 pandemic, testing strategies focused mainly on symptomatic individuals and individuals in high-risk groups (e.g., healthcare workers, staff and residents of long-term care facilities). The impact of these strategies on the reproduction number has been investigated in a modelling study [Grassly et al. (2020)]. Later into the pandemic, many countries expanded testing to include individuals, irrespective of whether they are displaying symptoms. Expanded testing strategies are based on the assumption that undetected asymptomatic and mild cases not reporting to healthcare can have a substantial impact on transmission of SARS-CoV-2. Population-wide testing at regular time intervals, also referred to as mass screening or universal testing, is now under serious consideration as a strategy to maintain incidence at manageable levels despite drastic reductions of control measures that are most disruptive to society [European Center for Disease Prevention and Control, (2020), Organisation for Economic Co-operation and Development (2020)]. Such an approach refers to testing a certain (substantial) part of the population, irrespective of the presence of symptoms, followed by isolation of infected subjects to interrupt transmission. The ultimate goal of such a strategy is to maintain the effective reproduction number *R*_*e*_ below 1, once it has been brought to such a level by the intensive measures. However, the effectiveness of such a strategy has not been determined. Apart from testing frequency, the effectiveness of regular universal testing depends on at least five other variables: (1) test sensitivity, which may vary according to the day since acquisition of infection [Kucirka et al. (2020)]; (2) the proportion of the population tested; (3) the effectiveness of isolation of infected persons; (4) the risk of transmission originating from subjects with false-negative test results and of (5) those that were not tested. Variables (4) and (5) relate to the level of reduction in *R*_*0*_ that is achieved by a basic set of control measures that remains in place. Subjects with false-negative test results may no longer adhere to these control measures and, therefore, may create conditions in which the number of secondary cases per primary case can grow above the threshold 1, and even approach levels observed at the onset of the pandemic, for which *R*_0_ is typically between 2 and 3 [Li et al. (2020); Park M et al. (2020)].

We use an epidemiological model to determine the effects of regular universal testing, as defined above, on the effective reproduction number *R*_*e*_ of SARS-CoV-2, for different fractions of the population being tested at different time intervals, and with different levels of transmission for tested and non-tested subjects. For simplicity, we define effectiveness as an overall reproduction number due to the random testing strategy (*R*_*rt*_) below 1 for the whole population with different levels of reopening society. The basic scenario is that all measures are lifted for tested subjects, and that non-tested subjects adhere to measures that control SARS-CoV-2 transmission so that *R*_*e*_ = 1.3. We also analyze scenarios where all subjects adhere to measures that control SARS-CoV-2 transmission to *R*_*e*_ *=* 1.3 or where all measures are lifted for all subjects. Complete reopening of society (i.e., lifting of all measures) allows SARS-CoV-2 transmission with *R*_*e*_ that is equal to *R*_*0*_ which we assume to be either 2.0 or 2.5.

## Methods

In this model we distinguish individuals who participate in regular universal screening (‘testing individuals’) and those who do not (‘non-testing individuals’). We assume that testing individuals participate in high-risk transmission events (i.e., where large crowds gather), and, thus, could generate more secondary infections than non-testing individuals, who are not permitted at these high-risk events. We assume that testing and non-testing individuals mix randomly during low-risk events, and that during high-risk events there is only transmission between testing individuals. Given a test-strategy — which is defined by a combination of the proportion of the population who tests regularly and the time interval between subsequent tests — we calculate the number of secondary cases of testing and non-testing individuals and how they are distributed over these two categories. Knowledge of these numbers allows us to determine the overall reproduction number *R*_*rt*_ for a given testing strategy.

In the model, the probability of onward transmission and the test sensitivity depend on the time since acquisition of infection. Both are described by discrete time distributions in time units of day. We assume a generation time distribution which has a Weibull-shape with shape parameter of 2.2826 and scale parameter of 5.665 [Ferretti et al. (2020)]. The discretized probability distribution between day *t –* 1 and day *t* is denoted by *g*(*t*). We further assume that a fraction *p* of the individuals in the population gets tested every *k* days for SARS-CoV-2 (testing individuals). The sensitivity of this test depends on the time since the tested individual got infected. This time-dependent sensitivity, *s*(*t)*, was based on the results of [Gremmels et al. (2020), Moeren van der et al. (2020)], (see Figure 1 and Supplementary Table S1). In our basic scenario we assume the same sensitivity for an antigen-based test and a PCR-test. As there is little data to accurately determine the sensitivity of a test to detect infectiousness, we also performed a sensitivity analysis in which assume that on every day since acquisition of the virus test sensitivity for detecting infectiousness was 80%.

**Figure 1.**
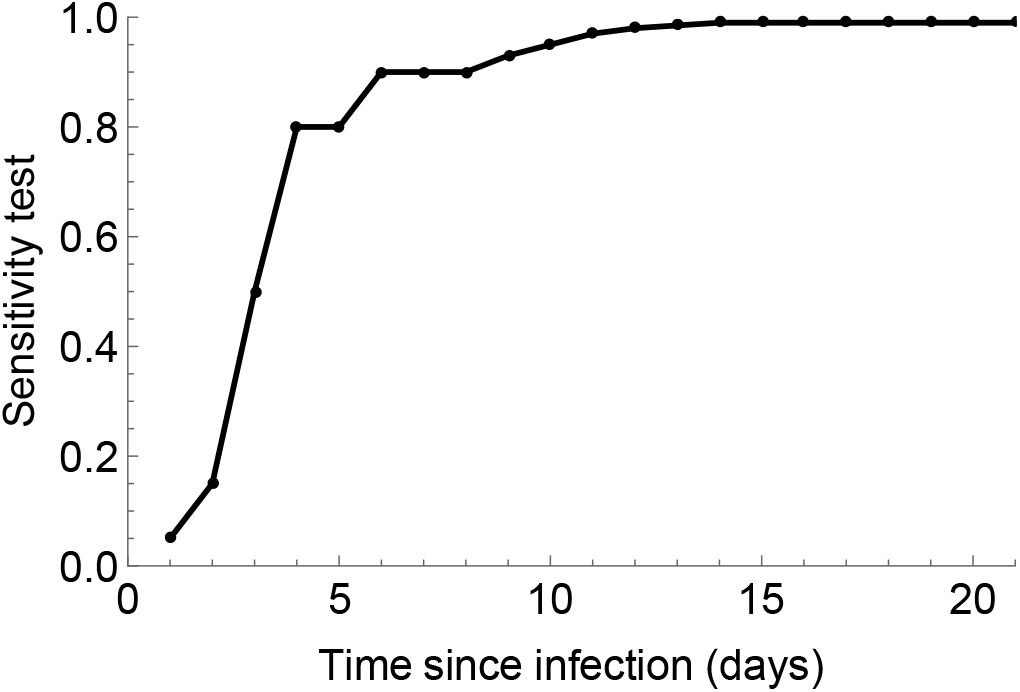
Sensitivity of the PCR-test as function of the time since acquisition given that the case is still infectious. See Table S1 for the data points.

We assume that non-testing individuals infect on average *R*_*n*_ individuals. This *R*_*n*_-value reflects the level of existing transmission prevention measures. We assume these infections to be proportionally distributed over testing and non-testing individuals, i.e., a non-testing individual infects, on average, *K*_*tn*_ *= pR*_*n*_ testing individuals and *K*_*nn*_ *=* (1 – *p*)*R*_*n*_ non-testing individuals (Figure 2). Testing individuals may have a higher reproduction number due to behavioral aspects, as having a negative test result may allow them to participate in higher-risk events. On the other hand, their reproduction number is reduced after a positive test result as the individual will then be isolated. A testing individual infects on average *K*_*nt*_ non-testing individuals and *K*_*tt*_ testing individuals (Figure 2). In the following we show how these numbers depend on the testing strategy.

**Figure 2.**
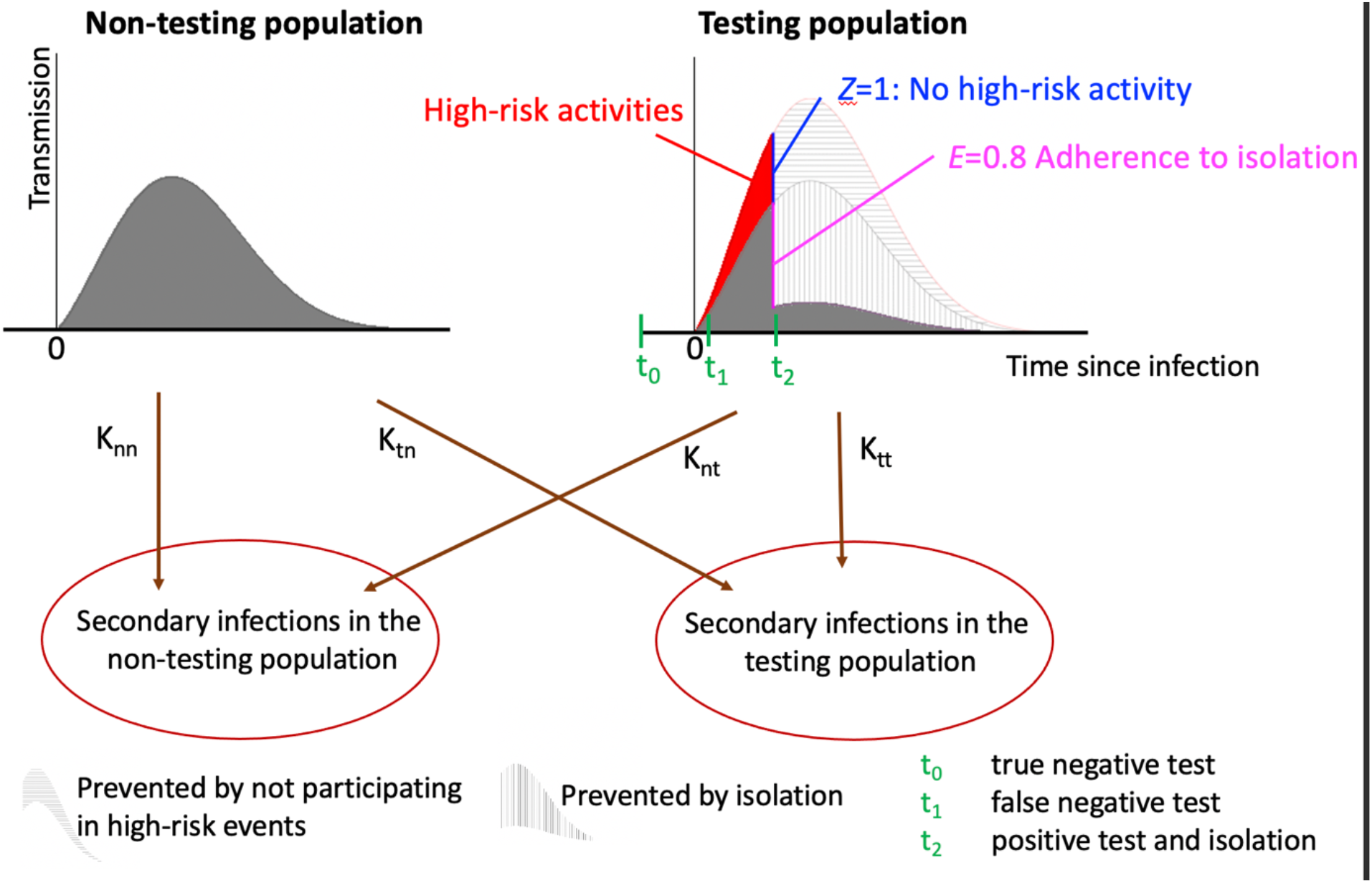
Schematic description of the model.

The probability *n*(*t*) for a testing individual to remain undetected at day *t* since infection, either because no test was performed or because all tests performed were false-negative, is given by

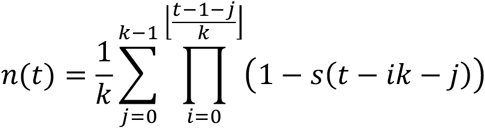

where we assumed that the risk to become infected is independent of the time since the last test and 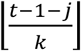 is the greatest integer that is less than or equal to 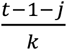 and *s*(*m*) *=* 0 if *m <* 1. The number of non-testing individuals infected by a testing individual (*K*_*nt*_) is given by

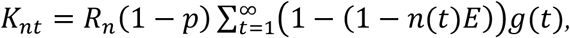

where the factor *E* represents the effectiveness of isolation on transmission prevention during low-risk events (*E =* 0 implies no effect, while *E =* 1 indicates full effectiveness). Our baseline value is *E =* 0.8, indicating that 80% of the transmissions in low-risk settings are prevented after an individual receives a positive test result.

The number of testing individuals infected by a testing individual (*K*_*tt*_) is given by

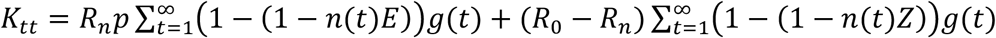

where *Z* denotes the effectiveness of preventing transmission during high-risk events after an individual receives a positive test result. Our baseline value of *Z* = 0 indicates that individuals with a positive test no longer participate in high-risk events.

The reproduction number *R*_*rt*_ with regular rapid testing of a fraction of the population is given by the largest eigenvalue of the next-generation matrix *K* [Diekmann et al. (2010)]

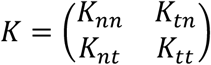

which is given by

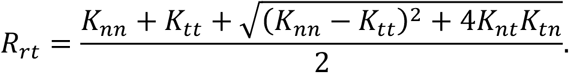

In summary, *R*_*rt*_ is the reproduction number in a population where regular rapid testing is implemented. All individuals in the tested population are tested at regular time intervals and are allowed to participate in high-risk events if tested negative. We analyzed how *R*_*rt*_ depends on the proportion of the population being tested and the time interval of testing. We assumed that this testing occurs in a population with all measures lifted (hence the potential for transmission resembles *R*_0_ = 2.5), or in a population where other measures have led to some sustained reduction of transmission (*R*_0_ = 2.0 or *R*_0_ = 1.3). We determine the requirements for regular testing intervals and coverage to control transmission in these three scenarios.

## Results

Based on the time-dependent sensitivity of diagnostic testing (Figure 1) the likelihood of infected subjects to remain undetected per day since acquisition of infection declines rapidly with more intensive testing (Figure 3). To detect an infected person around the time of highest infectivity (around day 5) with 50% probability, a screenings interval of 3 days or less is needed.

**Figure 3.**
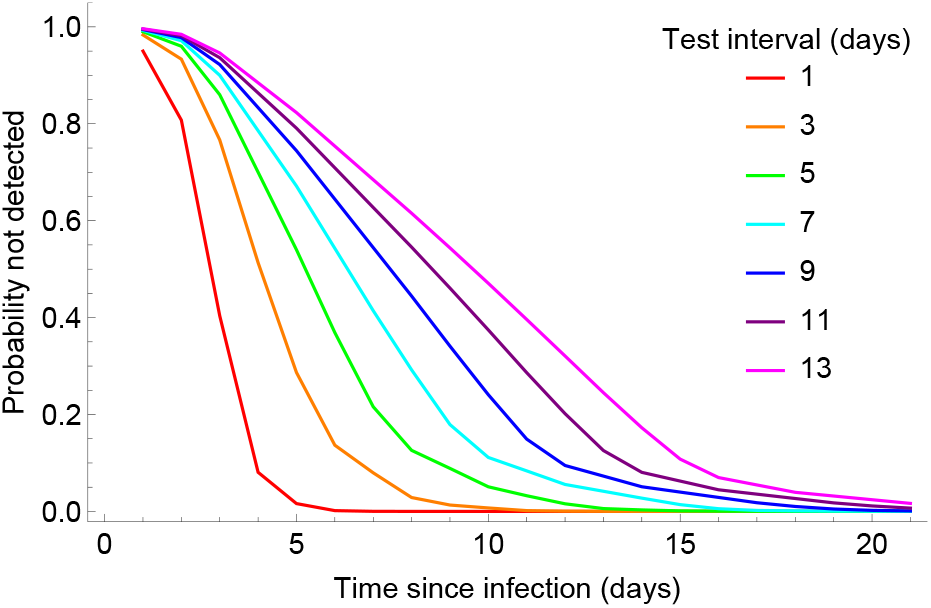
Probability as function of the time since infection for an infected individual to escape from being detected for varying times between two rapid tests. The test sensitivity equals that of the PCR-test.

In Figure 4 we have depicted scenarios in which the transmission level (*R*_*n*_) is identical for testing and non-testing individuals, i.e., testing individuals do not engage more in high-risk activities than non-testing individuals. If *R*_*n*_ is 1.5 — corresponding to a slight re-opening of society — at least 80% of the population needs to be tested every five days to bring a reproduction number of 1.5 to *R*_*rt*_ to a value below 1. In theory, regular testing can control an epidemic with a reproduction number of 3, but this would require daily testing of the whole population (Figure 4).

**Figure 4.**
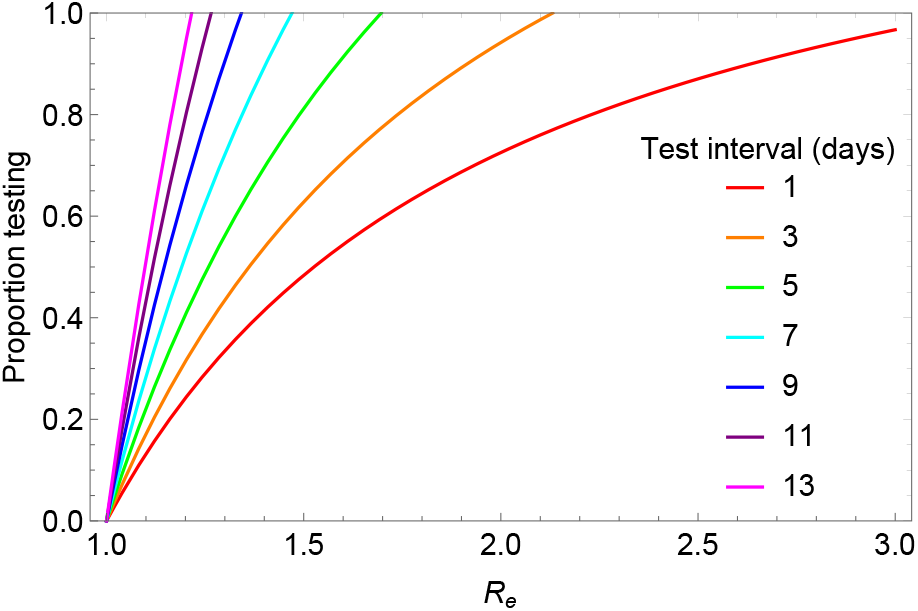
Lines depicting the critical values of the reproduction number achieved by a basic set of measures and the proportion of the population that gets tested in order to bring *R*_*rt*_ to one for different values of the time between tests. *E* = 0.8, there are no high-risk activities (*R*_*n*_ = *R*_*t*_) The test sensitivity equals that of the PCR-test.

Subsequently, in Figure 5 we have depicted the associations between test frequency, the proportion of the population being tested and the ‘risk behavior’ of those with false-negative test results for a test sensitivity as given in Table 1a. In these analyses we have assumed isolation efficacy of 80% and that all subjects tested positive will isolate and will refrain completely from high-risk activities. We assume that testing individuals without a positive test result (including those with a false-negative test result) will continue to participate in risk behavior that would lead to transmission of a virus with a corresponding expected number of new cases of 2.5, 2.0 or 1.3 (depending on the extent to which existing control measures have been lifted). The fraction of the population not tested has a risk behavior that would lead to transmission of a virus with a corresponding expected number of new cases of 1.3.

**Figure 5.**
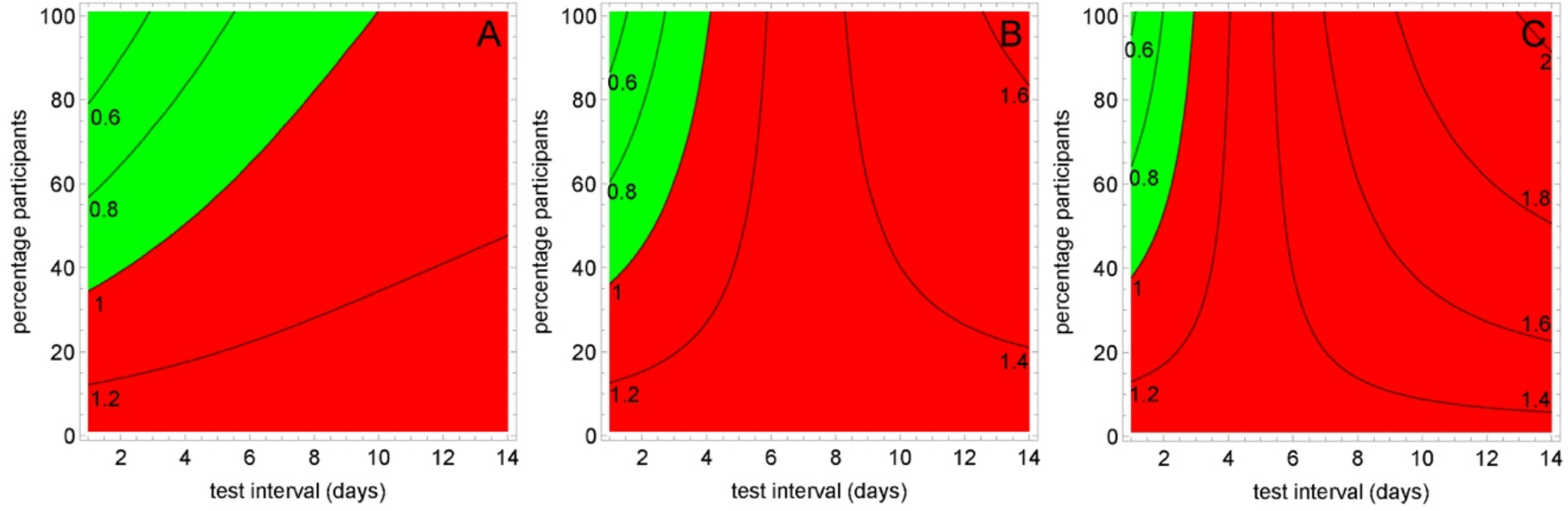
Reproduction number *R*_*rt*_ as a function of test intervals and percentage of participants. The contour lines define *R*_*rt*_, and green and red areas denote *R*_*rt*_ below and above 1, respectively. Time intervals between tests was between 1 and 14 days and percentage participants varied from 0% to 100% for an *R*_*n*_ of 1.3, a quarantine efficacy *E* of *0*.*8* and a total withdrawal from high-risk activities after a positive test (*Z* = 0). The reproduction number *R*_*t*_ was 1.3, 2.0 and 2.5 in panels A, B, and C, respectively. The test sensitivity equals that of the PCR-test.

**Figure 6.**
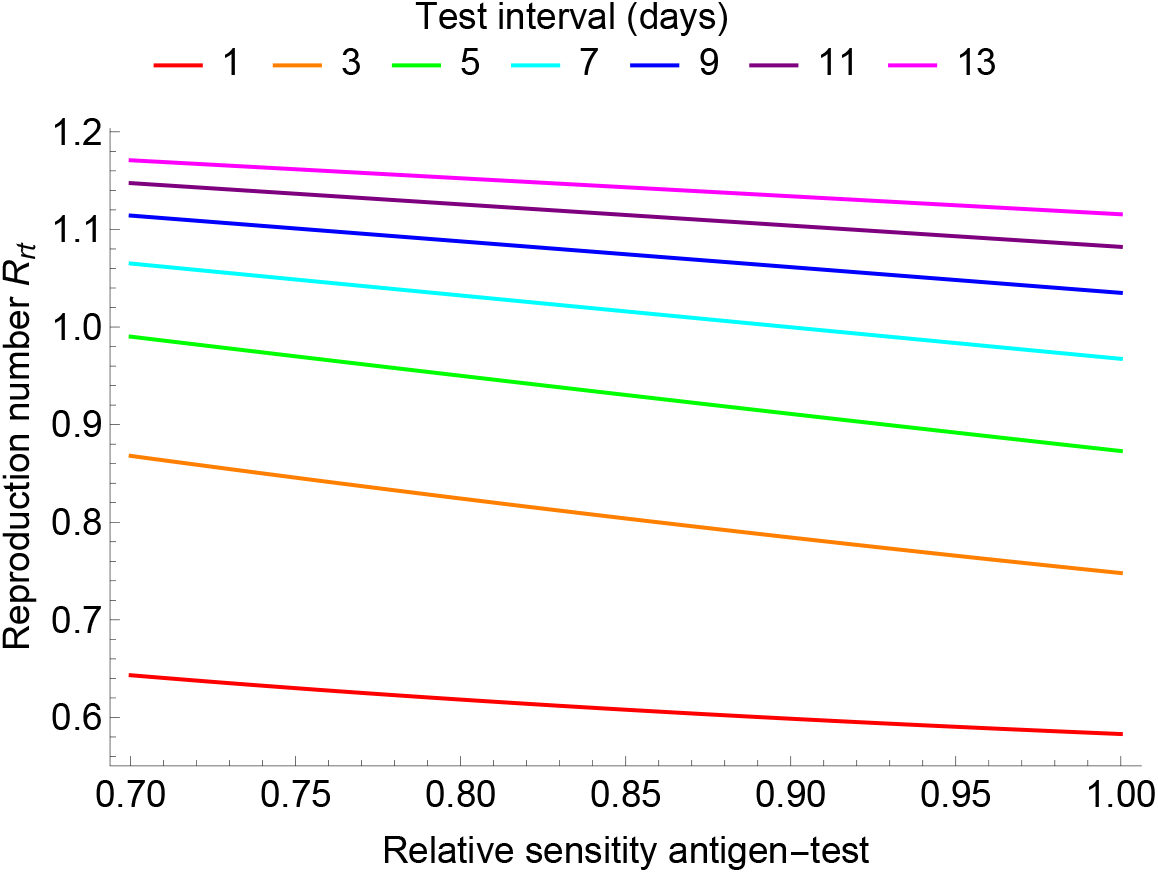
Dependence of the reproduction number *R*_*rt*_ on the sensitivity of the antigen test compared to the PCR-test. We used time intervals between tests of 1 to 13 days, 80% participation to testing, *R*_*n*_ = *R*_*t*_ of 1.3, and a quarantine efficacy *E=0*.*8*.

In Figure 5, the parameter region of testing frequency and the proportion being tested where the overall *R*_*rt*_ is below 1 is depicted in green. For *R*_*t*_ = 2.5 (Figure 5C) this would only be achieved if 100% of the population would be tested every three days, when using a rapid test method with similar sensitivity as PCR-based tests. For *R*_*t*_ = 1.3, testing 80% of the population once a week would be sufficient for controlling the epidemic (Figure 5Aa), for *R*_*t*_ = 2.0 testing 80% of the population every three days would be sufficient. With lower proportions tested, testing frequency should increase further to bring *R*_*rt*_ below 1. With a test that has a 10% lower sensitivity than the PCR test, the total population would need to be tested every 2.5 days to bring *R*_*rt*_ below 1 (Supplementary Figure 2).

A scenario with *R*_*n*_ = 1.3 reflects a situation with considerable control measures kept in place compared to the uncontrolled state where *R*_0_ = 2.5. In the Netherlands, this would reflect the situation in August 2020, with universal social distancing, working from home if possible, not working with (minor) respiratory tract infection symptoms, all large events prohibited, bars open only with seated placing, active testing with isolation of symptomatic people and contact tracing. In such circumstances, the proportion of the population to be tested weekly to reduce the overall *R*_*rt*_ to a value below 1 needs to be 60% when using PCR-based testing sensitivity and 80% for antigen-based testing sensitivity.

The impact of the test-sensitivity on the reproduction number is far less than the frequency of testing (Figure 7). Sensitivity analyses, in which we assume that on every day since acquisition of the virus test sensitivity for detecting infectiousness was 80% did not change interpretation (Supplementary Figures S3-S6).

## Discussion

The results of our analysis suggest that regular universal random screening followed by isolation of infectious individuals is not a viable strategy to reopen society after controlling a pandemic wave of SARS-CoV-2. The underlying mechanisms for failure are the suboptimal sensitivity of any testing method during the first days after acquisition in combination with an average generation time of four days and a basic reproduction number between 2 and 3.

To our knowledge, our study is the first to address the prospects of regular screening for SARS-CoV-2 infection irrespective of whether the individuals have symptoms for controlling the ongoing pandemic. Previous studies focused on the assessment of screening strategies for symptomatic individuals only and/or individuals in high-risk groups such as long-term care facilities, university and college students [Gassly et al. (2020), Delaunay et al. (2020), Paltiel et al (2020), Chang et al (2020), Martin et al. (2020)].

As any model, our model is a simplification of reality, but the major dynamical aspects of the transmission of SARS-CoV-2 have been included. Importantly, some parameters were deliberately rather optimistic, such as the effectiveness of isolation practices, the complete decline in high-risk activities for those being tested positive, absence of introductions from abroad and equal test sensitivity for antigen and PCR-based tests. Our findings are, therefore, most likely too optimistic.

The underlying mechanism leading to failure is the inevitable return, when lifting all previously imposed distancing measures, of transmission to values approaching *R*_0_, as long as herd immunity is too low to substantially reduce the effective reproduction number by the natural infection process. In the Netherlands, as in many other countries, an estimated 5-10% of the population had detectable immunity against SARS-CoV-2 after the first pandemic wave [National Institute for Public Health and the Environment, 2020].

Simplifications of our model are the absence of explicit age or network structure and a uniform view of human behavior that only differentiates between testing and non-testing individuals. Our quantitative results should, therefore, be interpreted with care. Younger individuals have a lower risk of complications due to COVID-19 and may, therefore, be more likely to participate in higher-risk activities and will have — on average — higher numbers of contacts. This could lead to higher reproduction numbers for testing individuals, making our estimates too optimistic. We did also not consider other factors important for implementation of regular universal random testing, such as costs, logistics, technical feasibility, resource availability and barriers to testing.

Naturally, scenarios must be developed that allow society to reopen safely after controlling a pandemic wave. Here, we have analyzed the likelihood of repeated universal random screening being successful, as this strategy is now proposed in many countries. Other population-wide testing approaches that have been used in various countries include household testing [David et al. (2020), Park Y et al. (2020)] and testing of incoming travelers [US Centers for Disease Control and Prevention (2020), Normile et al. (2020)]. Further analytical work will include scenarios with augmented contact tracing and screening based on observed transmission patterns, occurrence of high-risk events, or targeting high-risk populations, and combinations of these. Contact tracing may have different effects on the effectiveness of universal screening strategies. For instance, contact tracing may only substantially add to universal screening effectiveness if contacts are traced faster than through regular universal screening. On the other hand, if contact tracing occurs too fast, test sensitivity may be low and infectiousness may be missed if not rescreened after a few days. This can be overcome by quarantining traced contacts, even with negative test results.

The sensitivity to detect infectious subjects in the early stage of infection is an important parameter in our model. Yet, in the absence of accurate data, the infectiousness in time can only be derived indirectly. The relation between test-sensitivity, infectiousness and the effect of isolation is complex, for at least two reasons. First, the generation distribution describes the average proportion of infected subjects being infectious since the moment of virus acquisition (Ferretti et al. 2020). This distribution depends on preventive measures taken. For instance, effective isolation after symptom onset, will reduce the likelihood of transmission at the later stages of the infectious period, thereby changing the generation time and shortening the generation interval. The correct distribution of infectiousness, therefore, depends on the measures taken to reduce the reproduction number.

Second, there is considerable variation in timing of infectivity between individuals. As infectivity, viral load and test sensitivity are correlated, this implies similar variation in test-sensitivity between individuals. Subjects being highly infectious shortly after virus acquisition will most likely have high viral loads at that time, and hence, higher likelihoods to be detected by testing. Consider a test that will detect all individuals who are infectious, but not those in their latency period, and all detected subjects will be isolated immediately with 100% effective isolation. If 10% of infected individuals are infectious one day after acquisition, only 10% of all individuals tested on that day, respective to acquisition, will be detected and isolated. Yet, this will prevent transmission at that point of time and only latently infected individuals that become infectious at later stages will contribute to transmission. Assuming a test sensitivity for detecting infectious subjects of 10% will then be an underestimate, whereas assuming a test sensitivity of 100% will be an overestimate. We have, therefore, considered several scenarios for the infectiousness profile and test sensitivity.

We urge policy makers to carefully determine the likelihood of success of future strategies before widespread implementation. Mathematical modelling is a possible tool for such analyses, that may also guide targeted experiments to validate model findings or to obtain more quantitative information on critical parameters.

In conclusion, our findings provide evidence that a strategy of regular universal random testing would require unrealistic high testing frequencies to reopen society while maintaining control of virus transmission.

## Data Availability

The Mathematica code can be obtained from the authors.

## Supplementary Figures

**Figure S1.**
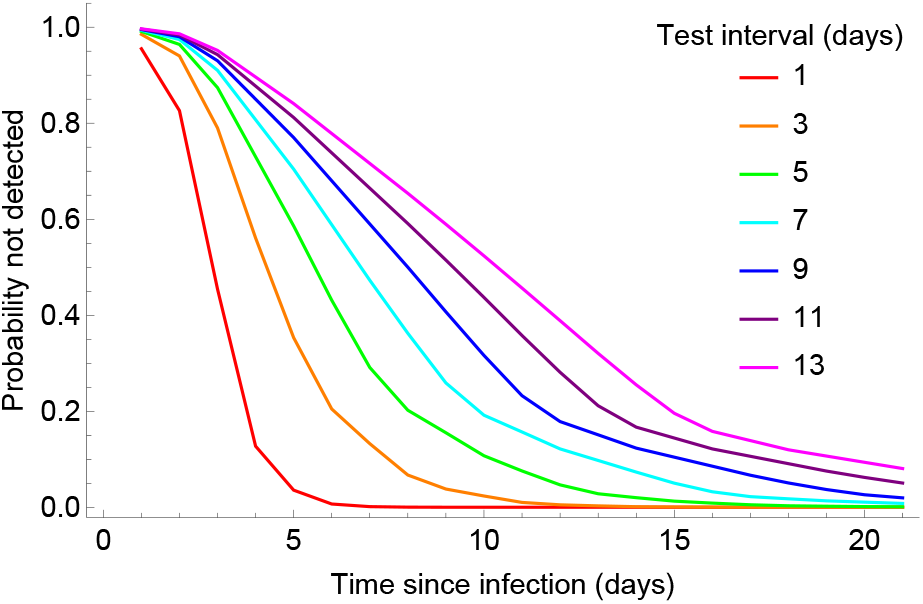
Probability as function of the time since infection for an infected individual to escape from being detected for varying times between two rapid tests. We assume the sensitivity of the antigen test to be 10% lower than the sensitivity of the PCR-test (Table S1).

**Figure S2.**
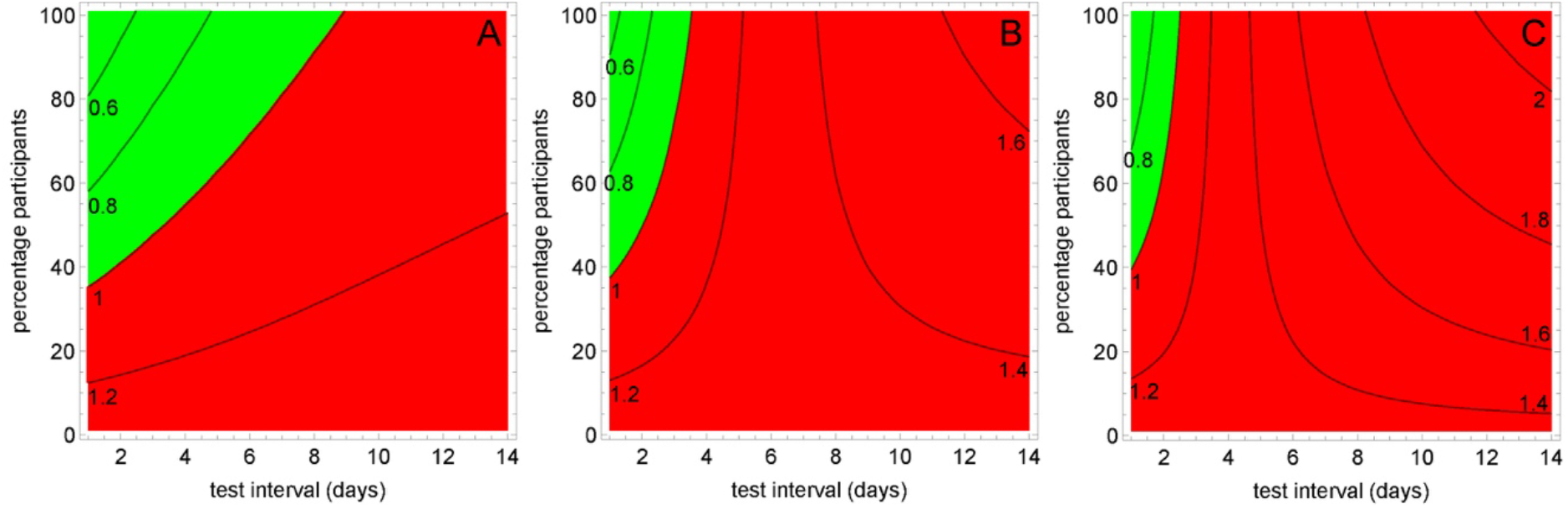
Reproduction number *R*_*rt*_ as a function of test intervals and percentage of participants. The contour lines define *R*_*rt*_, and green and red areas denote *R*_*rt*_ below and above 1, respectively. Time intervals between tests was between 1 and 14 days and percentage participants varied from 0% to 100% for an *R*_*n*_ of 1.3, a quarantine efficacy *E* of *0*.*8* and a total withdrawal from high-risk activities after a positive test (*Z* = 0). The reproduction number *R*_*t*_ was 1.3, 2.0 and 2.5 in panels A, B, and C, respectively. We assumed the sensitivity of the rapid antigen test to be 10% lower than the sensitivity of the PCR-test (Table S1).

**Figure S3.**
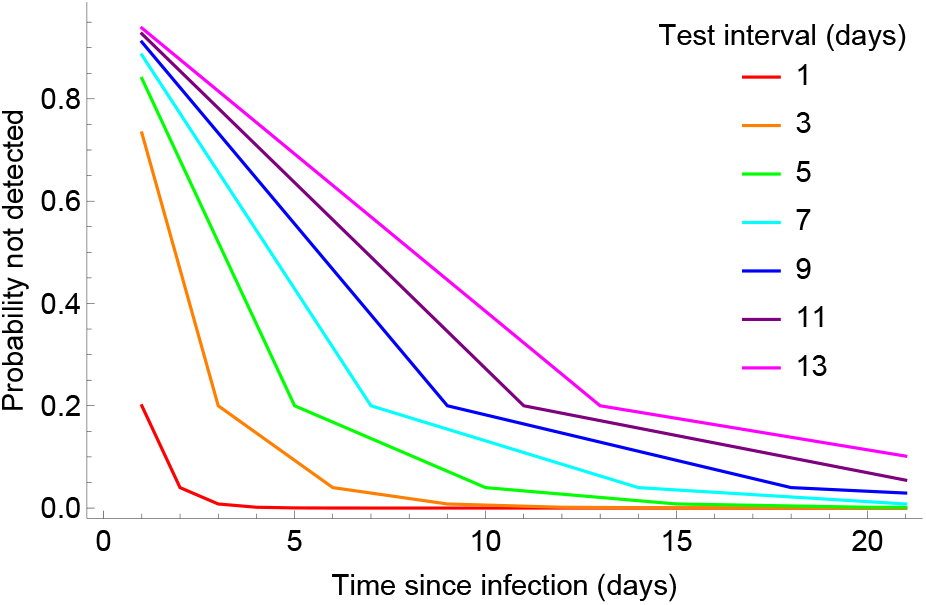
Probability as function of the time since infection for an infected individual to escape from being detected for varying times between two rapid tests. The test sensitivity is 80%, independent of the time since acquisition.

**Figure S4.**
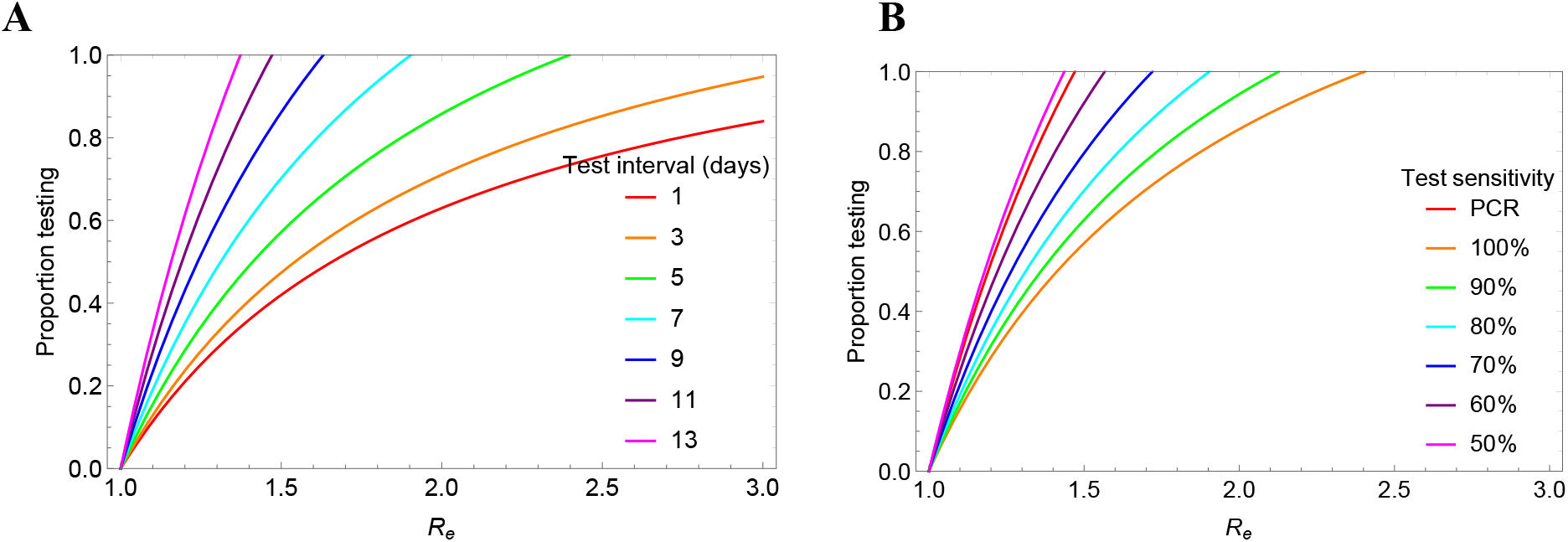
Lines depicting the critical values of the reproduction number achieved by a basic set of measures and the proportion of the population that gets tested in order to bring *R*_*rt*_ to one for different values of the time between tests. *E* = 0.8, there are no high-risk activities (*R*_*n*_ = *R*_*t*_) Panel A: The test sensitivity is 80%, independent of the time since acquisition. Panel B: Weekly screening with either the test sensitivity of Table S1 (PCR) or a test with a sensitivity independent of the time since acquisition.

**Figure S5.**
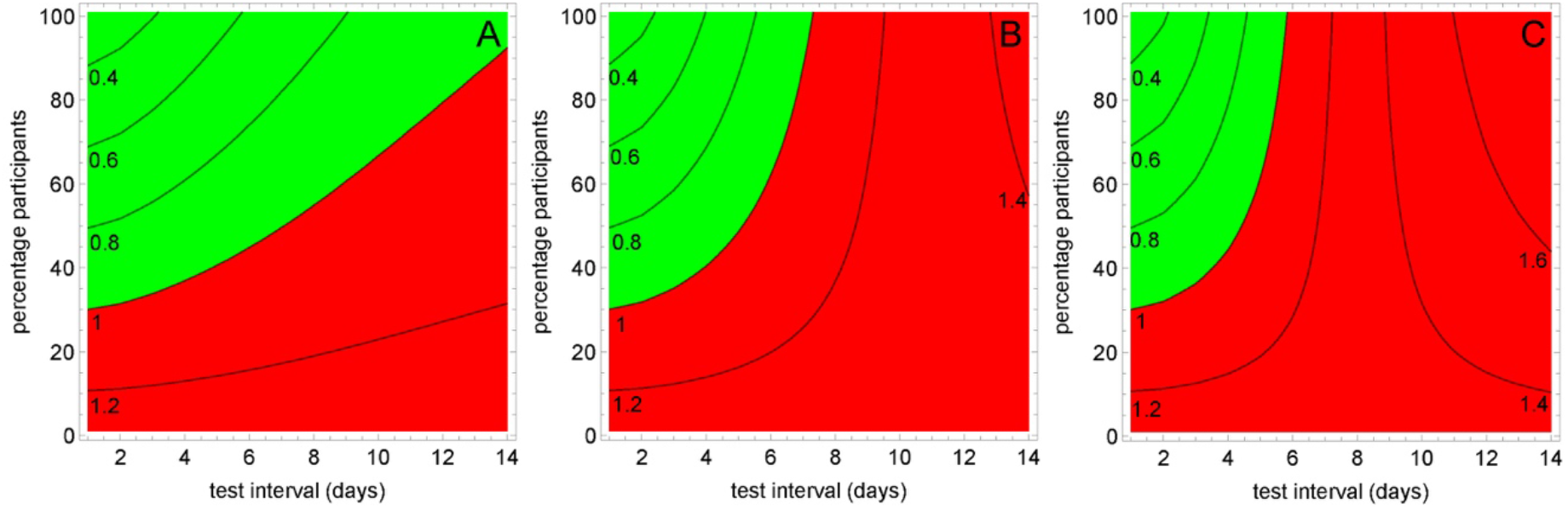
Reproduction number *R*_*rt*_ as a function of test intervals and percentage of participants. Time intervals between tests was between 1 and 14 days and percentage participants varied from 0% to 100% for an *R*_*n*_ of 1.3, a quarantine efficacy *E* of *0*.*8* and a total withdrawal from high-risk activities after a positive test (*Z* = 0). The reproduction number *R*_*t*_ was 1.3, 2.0 and 2.5 in panels A, B, and C, respectively. The test sensitivity equals 80%, irrespective of the time since acquisition.

**Figure S6.**
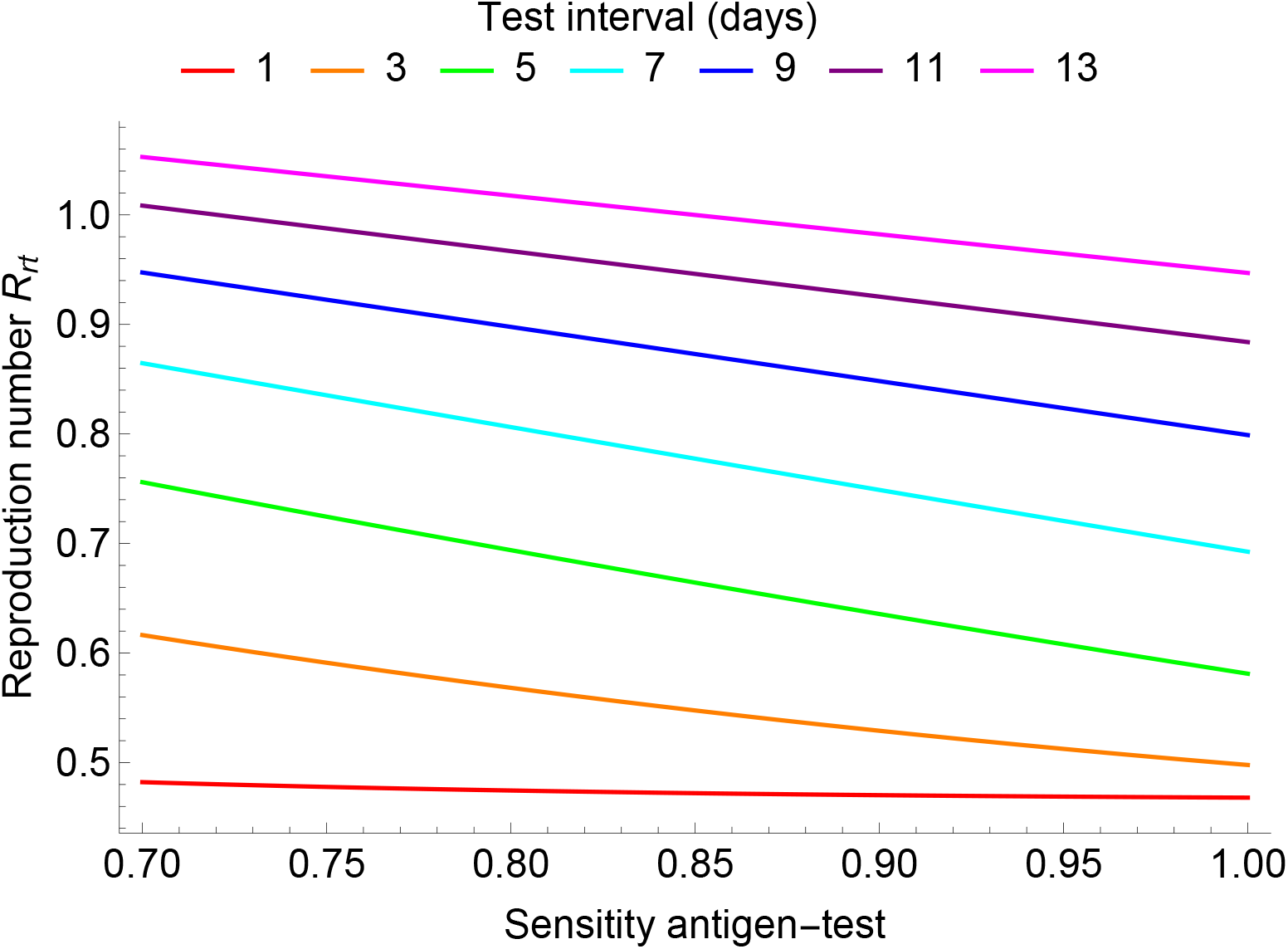
Dependence of the reproduction number *R*_*rt*_ on the sensitivity of the antigen test. We used time intervals between tests of 1 to 13 days, 80% participation to testing, *R*_*n*_ = *R*_*t*_ of 1.3, and a quarantine efficacy *E=0*.*8*. The test sensitivity of the antigen test is assumed to be independent of the time since acquisition.

## Supplementary Table

**Table S1:** Sensitivity test of the PCR-test and the values if the rapid antigen-test is 10% less sensitive than the PCR-test.

| Day | 1 | 2 | 3 | 4 | 5 | 6 | 7 | 8 | 9 | 10 | 11 | 12 | 13 | >13 |
| --- | --- | --- | --- | --- | --- | --- | --- | --- | --- | --- | --- | --- | --- | --- |
| Sens. PCR (%) | 5 | 15 | 5 | 80 | 90 | 90 | 90 | 90 | 93 | 95 | 97 | 98 | 98.5 | 99 |
| Sens. antigen test (%) | 4.5 | 13.5 | 45 | 72 | 81 | 81 | 81 | 81 | 83.7 | 85.5 | 87.3 | 88.2 | 88.65 | 89.1 |

